# Corporal punishment acceptability among university students without childhood maltreatment experiences: Associated factors and subfactor-specific patterns

**DOI:** 10.64898/2026.11.18.26368169

**Authors:** Ayano Kit, Asuka Miyazaki, Naomi Kitano, Tetsuji Okawachi, Andari Wuri Astuti, Sumihisa Honda

**Author notes:** **Corresponding author:**Ayano Kit, MPH, PhD. Department of Health Sciences, Kobe University Graduate School of Medicine, Hyogo, Japan Address: 7-10-2 Tomogaoka, Suma-ku, Kobe, Hyogo 654-0142, Japan (Asuka Miyazaki).

## Abstract

**Background:** Acceptance of corporal punishment and other harmful behaviors toward children may persist even among young adults without childhood maltreatment experiences.

**Objective:** To identify factors associated with the acceptability of corporal punishment and related harmful behaviors toward children among university students without childhood maltreatment experiences, and to examine whether these associations differ across types of harmful behavior.

**Participants and Setting:** An online cross-sectional survey was conducted at four Japanese universities in June 2024. Of 699 students analyzed, 568 reported no childhood maltreatment experiences.

**Methods:** Participants rated the acceptability of 22 behaviors as discipline. Exploratory factor analysis identified three subfactors: neglect and psychological maltreatment, harsh physically aggressive behaviors, and less severe aggressive behaviors. Multivariable linear regression analyses were conducted among students without childhood maltreatment experiences. Higher scores indicated lower acceptance.

**Results:** More egalitarian gender-role attitudes (B = 1.40, p = .001) and less frequent school-based physical violence experiences (B = 1.49, p = .024) were associated with lower overall acceptance of corporal punishment and related harmful behaviors. Associated factors differed across subfactors. Witnessing psychological violence, learning about child rearing, and enrollment in health, welfare, or rehabilitation-related faculties were associated with lower acceptance of less severe aggressive behaviors.

**Conclusions:** Factors associated with acceptability differed across types of harmful behavior, even among students without childhood maltreatment experiences. These findings suggest the importance of prevention education addressing gender-role attitudes, recognition of harmful disciplinary practices, and child development. Educational opportunities throughout adolescence and young adulthood may help promote attitudes that reject violence against children.

## 1. Introduction

### 1.1. Corporal punishment in the prevention of child maltreatment

Child maltreatment, including physical abuse, psychological abuse, sexual abuse, and neglect, is an important public health issue that has long-term consequences for children’s physical and psychological health and development. Its prevention requires not only responses after maltreatment has occurred but also a primary prevention approach that promotes social norms and values that reject violence (Lilleston et al., 2017; World Health Organization, 2026). The WHO defines corporal punishment as punishment in which physical force is used and intended to cause some degree of pain or discomfort, however light (World Health Organization, 2025). Importantly, corporal punishment and discipline are not synonymous. Discipline refers to caregiving practices that support children in developing behaviors and self-regulation skills necessary for social life and does not require the infliction of physical pain. Therefore, corporal punishment cannot be justified as a form of discipline and should not be considered acceptable. In Japan, a 2019 legal amendment prohibited parents and other persons exercising parental authority from using corporal punishment when disciplining children, and the prohibition came into effect in April 2020 (Ministry of Health, Labour and Welfare, 2019). However, the legal prohibition of corporal punishment does not necessarily eliminate its social acceptance; efforts to address perceptions of corporal punishment and related social norms are also needed (Lilleston et al., 2017; Zolotor & Puzia, 2010). Furthermore, corporal punishment does not necessarily occur in isolation and may coexist with psychological aggression and other harmful child-rearing practices. A systematic comparison of measures of harsh parenting and child maltreatment found substantial overlap in the behaviors assessed by the two constructs, including not only physical behaviors but also psychologically harmful behaviors such as threatening and yelling (Backhaus et al., 2023). Physical punishment has also been associated not only with physical abuse but with other forms of child maltreatment, including psychological maltreatment and neglect, and may escalate into more severe maltreatment (Afifi et al., 2017). In severe cases, escalating harsh physical punishment may result in serious injury or death (Wilson et al., 2023). Thus, focusing on the acceptance of corporal punishment, psychological aggression, and other harmful behaviors as forms of “discipline” is important for preventing more severe violence against children and child maltreatment.

### 1.2. Formation of attitudes toward corporal punishment

Attitudes toward corporal punishment are unlikely to emerge suddenly when an individual becomes a parent; rather, they may develop through various experiences and learning processes from childhood through adolescence. Childhood experiences of corporal punishment have frequently been associated with later support for corporal punishment, and a recent scoping review found that the majority of studies supported the intergenerational transmission of corporal punishment (Xu et al., 2026). However, this association is not uniform, and attitudes toward corporal punishment may differ depending on the type of violence experienced and how those experiences are perceived (Bell & Romano, 2012; Gagné et al., 2007; Wadji et al., 2023).

At the same time, young people’s acceptance of corporal punishment may not be shaped solely by parenting experiences within the family. During development, individuals are exposed to a range of experiences outside the home, including experiencing or witnessing violence at school or in the community, interactions with adults and peers, and opportunities to learn about children and parenting. Indeed, research among adolescents has shown that direct exposure to and witnessing of violence are associated with subsequent attitudes toward violence (Murshid & Murshid, 2019). Through such experiences, young people may learn which behaviors are considered acceptable, while perceived social norms—such as the extent to which family members, friends, and the wider community are believed to accept corporal punishment—may also contribute to the formation of their own attitudes toward corporal punishment (Lilleston et al., 2017; Taylor et al., 2011).

Educational and professional learning experiences may also be associated with perceptions of corporal punishment. Among Japanese university students, childhood experiences of corporal punishment were associated with current endorsement of corporal punishment, whereas students majoring in nursery education showed lower levels of endorsement (Kitano et al., 2018). Therefore, understanding the formation of attitudes toward corporal punishment among young people requires consideration not only of their own experiences of corporal punishment or child maltreatment but also of violence-related experiences outside the home, interactions with others, educational and learning experiences, and the broader social values surrounding these experiences.

### 1.3. Opportunities for prevention among young adults without childhood maltreatment experiences

From the perspective of child maltreatment prevention, it is important to examine not only individuals with childhood maltreatment experiences but also how factors other than such experiences may contribute to the formation of attitudes toward corporal punishment. Wadji et al. (2023) found that associations between childhood maltreatment experiences and the perceived acceptability of maltreatment differed according to maltreatment type. In particular, no clear associations were found for physical abuse, emotional maltreatment, or exposure to domestic violence, highlighting the need to consider factors beyond individuals’ own maltreatment experiences. Accordingly, among young adults without childhood maltreatment experiences, examining how experiences during development, interactions with others, learning opportunities, and social norms and values are associated with attitudes toward corporal punishment may help identify potentially modifiable factors involved in the formation of these attitudes.

This perspective also has important implications for child maltreatment prevention. Child maltreatment is often difficult to detect from outside the family, and only a proportion of children who experience maltreatment come to the attention of formal support services (World Health Organization, 2026). Prevention efforts should therefore extend beyond determining how to intervene in families in which maltreatment has already occurred. Schools, universities, communities, and professional education settings may provide opportunities for more upstream prevention by promoting attitudes that reject corporal punishment before young people later interact with children through their professional roles or as parents.

In this context, university students constitute an important population for study. Adolescence and young adulthood are periods in which individuals have already accumulated experiences within their families, schools, and communities and have developed relatively established values, while many have not yet entered the stage of raising their own children. A study of university students found that more favorable attitudes toward corporal punishment were associated with a greater intention to use corporal punishment with their own future children and highlighted the potential for educational interventions in university settings (Policastro et al., 2024). In addition, university students who reported having been spanked during childhood were more likely to express favorable attitudes toward the use of spanking as a disciplinary practice (Love et al., 2025). These findings suggest that individual differences in attitudes toward corporal punishment are already observable during the university years and may be related to both prior experiences and intentions regarding future use of corporal punishment. Moreover, universities also educate not only young people who may become parents in the future but also future professionals in health care, welfare, education, and other fields that involve working with children and families. Identifying factors associated with the acceptance of corporal punishment among university students may therefore provide a basis for developing preventive strategies through a range of educational opportunities, including professional education, as well as for preventing future violence against children within families.

Perceptions of corporal punishment and other harmful behaviors toward children may also differ according to the type and severity of the behavior. Previous studies have shown that both the perceived acceptability of child maltreatment and its associated factors vary across maltreatment types (Wadji et al., 2023). More overt and severe abusive behaviors tend to be more readily recognized as maltreatment, whereas psychological maltreatment and neglect may be less readily recognized (Lui et al., 2022; Morgan et al., 2026). Therefore, rather than treating the acceptance of corporal punishment and related harmful child-rearing practices as a single, undifferentiated attitude, it is necessary to consider differences in the nature and severity of these behaviors when examining associations between their acceptability and potential background factors.

### 1.4. The present study

Accordingly, this study aimed to assess the acceptability of corporal punishment and related harmful behaviors toward children among Japanese university students, with a particular focus on students without childhood maltreatment experiences. Specifically, the study had two objectives: first, to explore factors associated with the acceptability of corporal punishment and related harmful behaviors toward children, focusing on developmental experiences, learning opportunities, and social and cultural factors; and second, to examine whether the factors associated with acceptability differed according to the nature of the harmful behaviors toward children.

We hypothesized, first, that even among young people without childhood maltreatment experiences, the acceptability of corporal punishment and related harmful behaviors would be associated with various experiences and learning opportunities within and outside the family, interactions with others, and social values. Second, we hypothesized that the factors associated with acceptability would differ according to the nature of the harmful behaviors toward children.

By addressing these objectives, this study sought to provide a basis for identifying potentially modifiable factors associated with the acceptability of corporal punishment and for informing primary prevention strategies aimed at reducing violence against children and child maltreatment.

## 2. Methods

### 2.1. Participants and recruitment

This cross-sectional study was conducted from June 3 to 10, 2024, using an anonymous, self-administered web-based questionnaire. Eligible participants were undergraduate students in Japan who were under 30 years of age and whose native language was Japanese. Students were recruited from four universities and were enrolled in faculties including Medicine, Dentistry and Pharmacology; Health, Welfare, and Rehabilitation; Early Childhood Education; and other fields (Nutrition, Sociology, and Engineering), spanning the first through sixth academic years. Study invitations were disseminated through classroom announcements and/or the universities’ internal networks (e.g., campus intranet and learning management systems). In total, 7,137 students were invited to participate.

### 2.2. Procedures, consent, and protection

Before accessing the questionnaire, students were required to read an online information sheet explaining the purpose of the study and procedures, including that the survey addressed sensitive topics such as child abuse and corporal punishment. The questionnaire took approximately 10 minutes to complete. The information sheet stated that participation was voluntary, that students could withdraw at any time for any reason, and that choosing not to participate would not affect their evaluation by lecturers. It also noted that some questions might cause psychological discomfort; therefore, students could skip any item or discontinue the survey without penalty. Contact information for the university’s student counseling and support services was provided so that students could seek assistance if needed.

After reading the information sheet, students indicated their agreement by clicking an “I agree to participate” button. Submission of the completed questionnaire was regarded as informed consent. No inquiries or complaints, including those related to physical condition or other concerns, were received from participants during the survey period.

The survey was administered via SurveyMonkey (SurveyMonkey Inc.), a secure web-based platform. A progress indicator was displayed during completion, and respondents could navigate back to previous pages and revise their answers before final submission. To minimize respondent burden and preserve autonomy when answering sensitive questions, all items were set as optional; thus, the amount of missing data varied by variable.

Because the survey link was distributed via classroom announcements and university networks, the number of students who viewed the invitation link could not be determined; therefore, view and participation rates were not calculated. However, a response rate based on the number of students invited was calculated (i.e., the number of submitted questionnaires divided by 7,137 invited students).

No technical restrictions (e.g., IP address checks or cookies) were used to prevent multiple submissions. However, the information sheet explicitly asked students to complete the questionnaire only once. In addition, receipt of the incentive required entry of a mobile phone number, which was collected solely for incentive delivery and was handled by the e-gift card provider rather than the research team; these identifiers were not linked to survey responses and were deleted by the provider within a few weeks after distribution.

Participants who completed the questionnaire received an e-gift card worth ¥500 (approximately US $3.32). Ethical approval was obtained from an institutional ethics committee in Japan (No.: 23-15). The initial dataset comprised 820 submissions. Of these, 86 were incomplete, 8 indicated that they did not agree to participate, and 27 had missing data on the AACP items and/or sex. Accordingly, data from 699 participants were included in the final analyses.

To protect confidentiality, only survey responses were used for analysis, and findings are reported in aggregate form so that no individual participant can be identified. The dataset was stored on a password-protected USB device, with access restricted to the project leader. In accordance with the approved protocol and ethics committee regulations, the data will be retained for a limited period and then securely deleted.

### 2.3. Measures

The web-based questionnaire included items on participants’ basic demographic characteristics, childhood family environment, household economic status, and experiences during elementary, junior high, high school, and university. Sex was self-reported using the response categories female, male, and other. The questionnaire did not separately assess sex assigned at birth and gender identity. In addition, questions assessed values and beliefs shaped by past experiences, such as gender role attitudes and views on marriage and childrearing. Several study-specific items were also used to assess participants’ experiences with children, experiences of being taught that corporal punishment is necessary, beliefs about corporal punishment, interest in child abuse, and learning experiences related to child rearing and the value of life. The full wording of these items, their response options, and the coding used in the analyses are provided in Appendix A of the Supplementary Material.

#### 2.3.1. Attitudes towards the acceptability of corporal punishment (AACP)

We assessed attitudes toward the acceptability of corporal punishment using 22 child abuse– related behaviors examined by Lee and Yasuyama (2002) in their study of parents’ perceptions of the boundary between discipline and child abuse. The 22 items represented various forms of corporal punishment and other potentially harmful behaviors toward children, such as harsh verbal scolding, slapping the hands, and withholding bathing. Several items correspond conceptually to behaviors included in the Parent–Child Conflict Tactics Scales (Straus et al., 1998). The item content was informed by a 1991 survey conducted in Japan by the “Women’s Issues Study Group,” a survey conducted by the Centre for the Prevention of Child Abuse, and information on child abuse provided by the Japanese Ministry of Health, Labour and Welfare (Lee & Yasuyama, 2002).

In the present study, these 22 behaviors were used to assess university students’ attitudes toward the acceptability of corporal punishment and other potentially harmful behaviors toward children. Each item was rated on a four-point Likert-type scale reflecting the extent to which the respondent considered the behavior acceptable as discipline (1 = acceptable as discipline, 2 = somewhat acceptable as discipline, 3 = somewhat unacceptable as discipline, and 4 = unacceptable as discipline). Item scores were summed to create the Attitudes toward the Acceptability of Corporal Punishment (AACP) score (range: 22–88), with higher scores indicating lower acceptance of corporal punishment and other potentially harmful behaviors toward children as discipline.

#### 2.3.2. Child maltreatment experiences

Childhood maltreatment experiences were assessed using five items adapted from the Adverse Childhood Experiences (ACE) questionnaire (Felitti et al., 1998) and its Japanese version developed and validated by Tsuboi (2014). The Japanese version was subsequently used in a peer-reviewed study in Japan (Yamagishi et al., 2022). For the present study, five items were selected and their wording was simplified and partially modified to facilitate comprehension and response by university students while retaining the core content of the corresponding ACE domains (Appendix B in the Supplementary Material). The five selected items corresponded to physical abuse, psychological/emotional abuse, sexual abuse, exposure to domestic violence, and neglect. Participants were asked whether they had experienced each event before the age of 18 years (“yes” or “no”). Those who answered “yes” to at least one of these five items were classified as having experienced childhood maltreatment, whereas those who answered “no” to all items were classified as having no childhood maltreatment experience.

#### 2.3.3. School-based experiences of corporal punishment and verbal abuse

To assess experiences of corporal punishment and verbal abuse in educational settings, we developed four original items addressing both direct experiences and witnessing of such behaviors by teachers or other adults in schools, after-school classes, or extracurricular settings. Specifically, participants were asked whether, during elementary, junior high, or high school, (1) they had ever been physically punished (e.g., hit or kicked) by a teacher or other school-related person, (2) had ever been verbally abused or insulted, (3) had witnessed others being physically punished, and (4) had witnessed others being verbally abused. Each item was rated on a 3-point scale (“never,” “occasionally,” or “frequently”). These questions were not derived from any existing standardized measure; rather, they were developed through discussions among the research team based on educational contexts in Japan. A pilot test was conducted with university students to ensure clarity and acceptability of the wording, and minor revisions were made according to participants’ feedback before the final survey was administered.

#### 2.3.4. Gender role attitudes

Participants’ gender role attitudes were measured with a single item asking, “Do you agree or disagree with the statement that ‘A husband should work outside, and a wife should protect the home’?” This item was adapted from the Cabinet Office’s national public opinion survey on gender equality in Japan (Cabinet Office, 2023). Responses were rated on a 4-point Likert scale ranging from 1 (“strongly agree”) to 4 (“strongly disagree”).

### 2.4. Statistical analysis

Statistical analyses were conducted in several steps. First, descriptive statistics were used to summarize participants’ demographic characteristics and the distributions of all study variables. Next, exploratory factor analysis was performed on the items of the Attitudes toward the Acceptability of Corporal Punishment (AACP) scale using the principal factor method with varimax rotation to examine its underlying factor structure. Correlations among the extracted subfactors were then assessed.

Because the aim of this study was to identify factors associated with attitudes toward the acceptability of corporal punishment and other potentially harmful child-rearing behaviors among individuals without a history of childhood maltreatment, subsequent univariate and multivariable analyses were restricted to participants without childhood maltreatment experiences. Univariate analyses were conducted to examine associations between participant characteristics, including sex, and the AACP total score as well as each subfactor score. Sex was also included as a candidate variable in the multivariable analyses. Normality of the score distributions was then assessed using the Kolmogorov–Smirnov test and the Shapiro– Wilk test. As the score distributions did not meet the assumption of normality, the Mann– Whitney U test was used for comparisons between two groups and the Kruskal–Wallis test was used for comparisons among three or more groups.

Based on the univariate results, variables with p-values < .30 were selected as candidates so as to avoid prematurely excluding potentially relevant variables in this exploratory analysis. In addition, variables considered relevant on the basis of prior literature and the study aims, including experiences of corporal punishment or witnessing violence in educational settings, were also considered for inclusion. Multiple linear regression analyses were then performed separately for the AACP total score and each subfactor score using a backward elimination procedure, with a removal criterion of p ≥ .10. Variance inflation factors were examined, and no serious multicollinearity was detected. However, because the purpose of this study was to explore broader background factors associated with attitudes toward corporal punishment, variables conceptually very close to the outcome itself were interpreted cautiously in model building. All analyses were conducted using SPSS version 25.0 for Windows (IBM, Tokyo, Japan), and statistical significance was set at p < .05.

## 3. Results

A total of 726 students completed the questionnaire, yielding a response rate of 10.2% based on the 7,137 invited students. Of these, 27 participants with missing data on the dependent variable and/or sex were excluded, and the remaining 699 participants were included in the analysis. Information on abuse experience was missing for eight participants; therefore, comparisons by abuse experience were conducted for 691 participants. Among them, 123 (17.6%) reported a history of abuse, whereas 568 (81.3%) did not. The mean age of the total sample was 20.2 years (SD = 1.89). Participants with abuse experiences were slightly older than those without abuse experiences (20.7 [SD = 2.09] vs. 20.1 [SD = 1.83], p = .004). In the total sample, most participants were female (72.1%), and the largest proportions were first-year students (29.6%) and fourth-year students (28.9%). Regarding place of birth, 44.9% were from the Kyushu area and 39.5% were from the Kansai area. Nearly half of the participants belonged to faculties related to medicine, dentistry, and pharmacology (45.1%), followed by health, welfare, and rehabilitation (36.3%).

Based on chi-square tests, significant differences between participants with and without abuse experiences were observed for perceived economic situation, current living status, and number of siblings. Participants with abuse experiences were more likely to report poorer economic circumstances, to live in other arrangements rather than alone or with parents, and to have fewer siblings (Table 1). Based on chi-square tests, significant differences between participants with and without abuse experiences were found in several variables related to experiences and values developed from elementary through high school (Table 2). Compared with those without abuse experiences, participants with abuse experiences were more likely to report experiences of physical violence toward themselves, psychological violence toward themselves, and psychological violence toward others during their elementary to high school years. They were also less likely to report interaction with the community, more likely to agree with discipline using corporal punishment, more likely to endorse traditional gender role views, and less likely to express a desire for marriage and for raising children.

**Table 1.** Demographic characteristics of the study participants and comparisons by abuse experience.

|  | Total<br>(N=699) |  | With abuse<br>experience<br>(n=123) |  | Without abuse<br>experience<br>(n=568) |  | *p-value* |
|  | n | (%) | n | (%) | n | (%) |  |
|---|---|---|---|---|---|---|---|
| Sex |  |  |  |  |  |  | .395 |
| Female | 504 | (72.1) | 84 | (68.3) | 412 | (72.5) |  |
| Male | 193 | (27.6) | 38 | (30.9) | 155 | (27.3) |  |
| Others | 2 | (0.3) | 1 | (0.8) | 1 | (0.2) |  |
| University grade |  |  |  |  |  |  | .715 |
| 1<sup>st</sup> | 207 | (29.6) | 31 | (25.2) | 174 | (30.6) |  |
| 2<sup>nd</sup> | 132 | (18.9) | 22 | (17.9) | 109 | (19.2) |  |
| 3<sup>rd</sup> | 96 | (13.7) | 17 | (13.8) | 75 | (13.2) |  |
| 4<sup>th</sup> | 202 | (28.9) | 42 | (34.1) | 159 | (28.0) |  |
| 5<sup>th</sup> | 38 | (5.4) | 6 | (4.9) | 32 | (5.6) |  |
| 6<sup>th</sup> | 22 | (3.1) | 5 | (4.1) | 17 | (3.0) |  |
| Unknown | 2 | (0.3) | 0 | (0.0) | 2 | (0.4) |  |
| Place of birth |  |  |  |  |  |  | .322 |
| Kanto area | 33 | (4.7) | 9 | (7.3) | 23 | (4.0) |  |
| Kansai area | 276 | (39.5) | 52 | (42.3) | 221 | (38.9) |  |
| Kyushu area | 314 | (44.9) | 49 | (39.8) | 261 | (46.0) |  |
| Others | 76 | (10.9) | 13 | (10.6) | 63 | (11.1) |  |
| Faculty |  |  |  |  |  |  | .142 |
| Medicine, Dentistry, and Pharmacology | 315 | (45.1) | 68 | (55.3) | 247 | (43.5) |  |
| Health, Welfare, and Rehabilitation | 254 | (36.3) | 36 | (29.3) | 214 | (37.7) |  |
| Child Education | 39 | (5.6) | 5 | (4.1) | 32 | (5.6) |  |
| Others | 76 | (10.9) | 13 | (10.6) | 61 | (10.7) |  |
| Unknown | 15 | (2.1) | 1 | (0.8) | 14 | (2.5) |  |
| Economic situation |  |  |  |  |  |  | .003 |
| Poor | 68 | (9.7) | 22 | (17.9) | 43 | (7.6) |  |
| Normal | 369 | (52.8) | 59 | (48.0) | 306 | (53.9) |  |
| Rich | 261 | (37.3) | 42 | (34.1) | 218 | (38.4) |  |
| Unknown | 1 | (0.1) | 0 | (0.0) | 1 | (0.2) |  |
| Current living status |  |  |  |  |  |  | .004 |
| Alone | 300 | (42.9) | 58 | (47.2) | 239 | (42.1) |  |
| With parents | 334 | (47.8) | 46 | (37.4) | 284 | (50.0) |  |
| Others | 63 | (9.0) | 19 | (15.4) | 43 | (7.6) |  |
| Unknown | 2 | (0.3) | 0 | (0.0) | 2 | (0.4) |  |
| Number of siblings （including the respondent） |  |  |  |  |  |  | .046 |
| 1 （only child） | 93 | (13.3) | 25 | (20.3) | 68 | (12.0) |  |
| 2 | 340 | (48.6) | 57 | (46.3) | 281 | (49.5) |  |
| ≥3 | 260 | (37.2) | 40 | (32.5) | 214 | (37.7) |  |
| Unknown | 6 | (0.9) | 1 | (0.8) | 5 | (0.9) |  |
| Living arrangement in childhood |  |  |  |  |  |  | .540 |
| With parents | 533 | (76.3) | 91 | (74.0) | 435 | (76.6) |  |
| Others | 166 | (23.7) | 32 | (26.0) | 133 | (23.4) |  |
| Partner status |  |  |  |  |  |  | .498 |
| Having | 227 | (32.5) | 43 | (35.0) | 180 | (31.7) |  |
| Not having | 470 | (67.2) | 80 | (65.0) | 386 | (68.0) |  |
| Unknown | 2 | (0.3) | 0 | (0.0) | 2 | (0.4) |  |
| Mother's final education |  |  |  |  |  |  | .200 |
| Under high school | 154 | (22.0) | 22 | (17.9) | 130 | (22.9) |  |
| Above high school | 516 | (73.8) | 97 | (78.9) | 413 | (72.7) |  |
| Others | 29 | (4.1) | 4 | (3.3) | 25 | (4.4) |  |
| Father's final education |  |  |  |  |  |  | .679 |
| Under high school | 160 | (22.9) | 25 | (20.3) | 129 | (22.7) |  |
| Above high school | 477 | (68.2) | 84 | (68.3) | 391 | (68.8) |  |
| Others | 61 | (8.7) | 14 | (11.4) | 47 | (8.3) |  |
| Unknown | 1 | (0.1) | 0 | (0.0) | 1 | (0.2) |  |

*Note.* The total analytic sample comprised 699 participants. Group comparisons by abuse experience were based on 691 participants because data on abuse experience were missing for eight participants. Group differences were examined using chi-square tests. Missing responses are shown as “Unknown.” For parental education, the “Others” category was excluded from the chi-square tests.

**Table 2.**
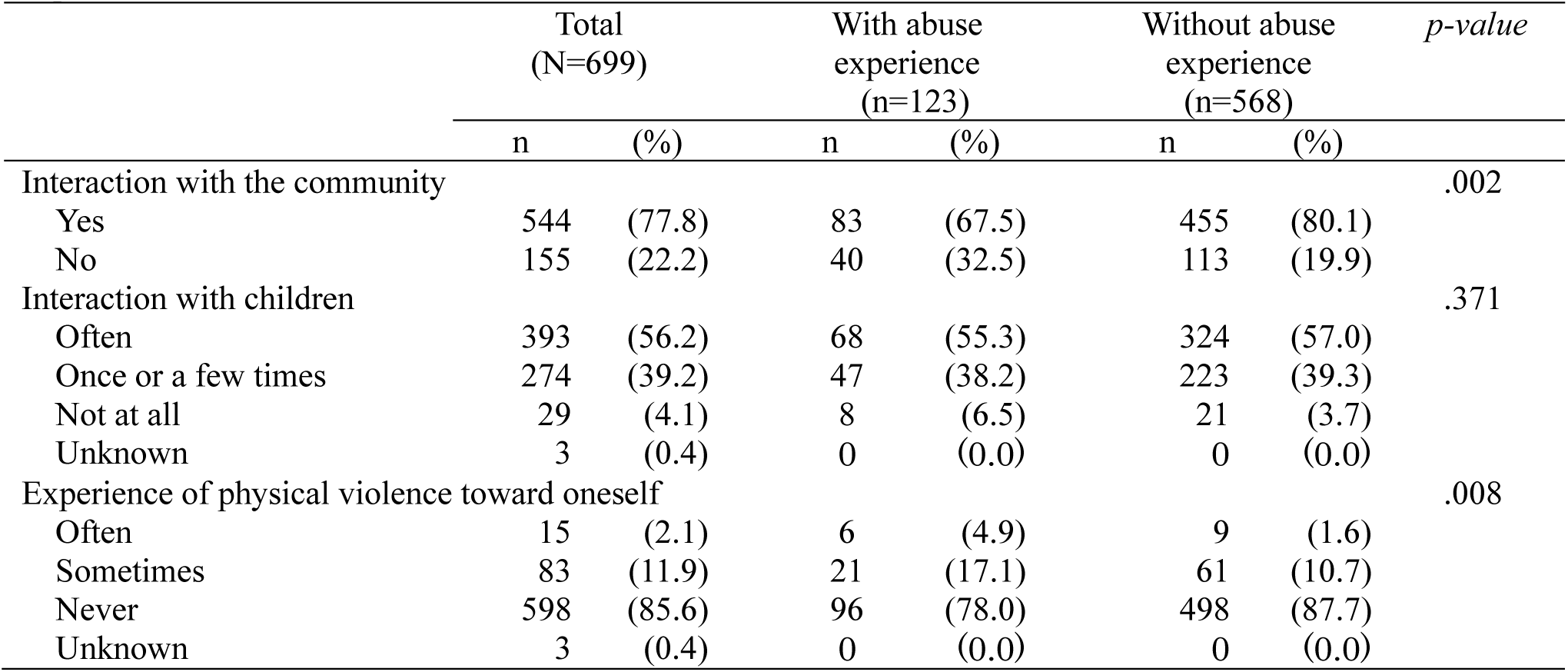
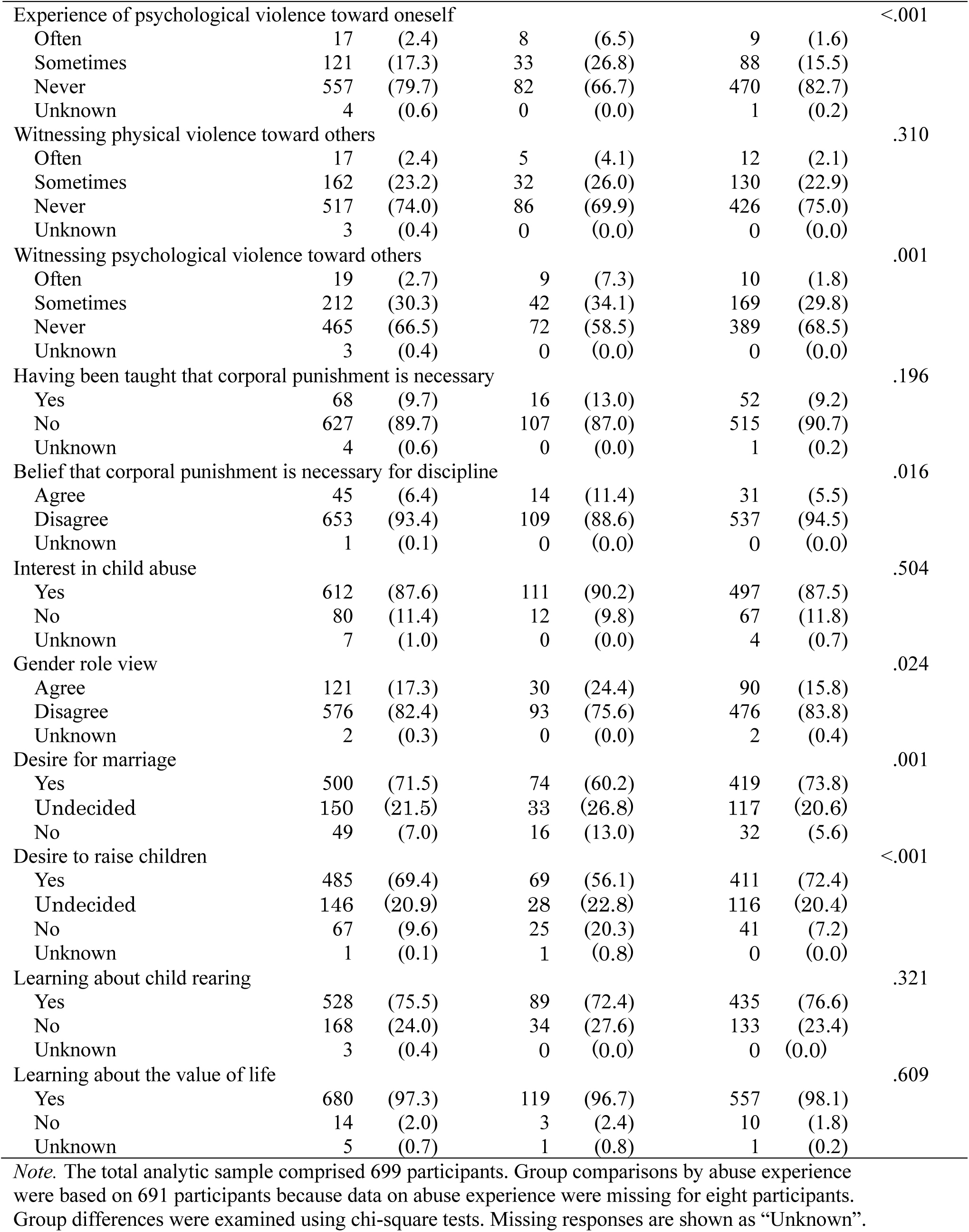
Experiences, beliefs, and learning opportunities among study participants and comparisons by abuse experience.

| Total<br>(N=699) |  | With abuse<br>experience<br>(n=123) |  | Without abuse<br>experience<br>(n=568) |  | *p-value* |
|---|---|---|---|---|---|---|---|
|  | n | (%) | n | (%) | n | (%) |  |
| Interaction with the community |  |  |  |  |  |  | .002 |
| Yes | 544 | (77.8) | 83 | (67.5) | 455 | (80.1) |  |
| No | 155 | (22.2) | 40 | (32.5) | 113 | (19.9) |  |
| Interaction with children |  |  |  |  |  |  | .371 |
| Often | 393 | (56.2) | 68 | (55.3) | 324 | (57.0) |  |
| Once or a few times | 274 | (39.2) | 47 | (38.2) | 223 | (39.3) |  |
| Not at all | 29 | (4.1) | 8 | (6.5) | 21 | (3.7) |  |
| Unknown | 3 | (0.4) | 0 | (0.0) | 0 | (0.0) |  |
| Experience of physical violence toward oneself |  |  |  |  |  |  | .008 |
| Often | 15 | (2.1) | 6 | (4.9) | 9 | (1.6) |  |
| Sometimes | 83 | (11.9) | 21 | (17.1) | 61 | (10.7) |  |
| Never | 598 | (85.6) | 96 | (78.0) | 498 | (87.7) |  |
| Unknown | 3 | (0.4) | 0 | (0.0) | 0 | (0.0) |  |
| Experience of psychological violence toward oneself |  |  |  |  |  |  | <.001 |
| Often | 17 | (2.4) | 8 | (6.5) | 9 | (1.6) |  |
| Sometimes | 121 | (17.3) | 33 | (26.8) | 88 | (15.5) |  |
| Never | 557 | (79.7) | 82 | (66.7) | 470 | (82.7) |  |
| Unknown | 4 | (0.6) | 0 | (0.0) | 1 | (0.2) |  |
| Witnessing physical violence toward others |  |  |  |  |  |  | .310 |
| Often | 17 | (2.4) | 5 | (4.1) | 12 | (2.1) |  |
| Sometimes | 162 | (23.2) | 32 | (26.0) | 130 | (22.9) |  |
| Never | 517 | (74.0) | 86 | (69.9) | 426 | (75.0) |  |
| Unknown | 3 | (0.4) | 0 | (0.0) | 0 | (0.0) |  |
| Witnessing psychological violence toward others |  |  |  |  |  |  | .001 |
| Often | 19 | (2.7) | 9 | (7.3) | 10 | (1.8) |  |
| Sometimes | 212 | (30.3) | 42 | (34.1) | 169 | (29.8) |  |
| Never | 465 | (66.5) | 72 | (58.5) | 389 | (68.5) |  |
| Unknown | 3 | (0.4) | 0 | (0.0) | 0 | (0.0) |  |
| Having been taught that corporal punishment is necessary |  |  |  |  |  |  | .196 |
| Yes | 68 | (9.7) | 16 | (13.0) | 52 | (9.2) |  |
| No | 627 | (89.7) | 107 | (87.0) | 515 | (90.7) |  |
| Unknown | 4 | (0.6) | 0 | (0.0) | 1 | (0.2) |  |
| Belief that corporal punishment is necessary for discipline |  |  |  |  |  |  | .016 |
| Agree | 45 | (6.4) | 14 | (11.4) | 31 | (5.5) |  |
| Disagree | 653 | (93.4) | 109 | (88.6) | 537 | (94.5) |  |
| Unknown | 1 | (0.1) | 0 | (0.0) | 0 | (0.0) |  |
| Interest in child abuse |  |  |  |  |  |  | .504 |
| Yes | 612 | (87.6) | 111 | (90.2) | 497 | (87.5) |  |
| No | 80 | (11.4) | 12 | (9.8) | 67 | (11.8) |  |
| Unknown | 7 | (1.0) | 0 | (0.0) | 4 | (0.7) |  |
| Gender role view |  |  |  |  |  |  | .024 |
| Agree | 121 | (17.3) | 30 | (24.4) | 90 | (15.8) |  |
| Disagree | 576 | (82.4) | 93 | (75.6) | 476 | (83.8) |  |
| Unknown | 2 | (0.3) | 0 | (0.0) | 2 | (0.4) |  |
| Desire for marriage |  |  |  |  |  |  | .001 |
| Yes | 500 | (71.5) | 74 | (60.2) | 419 | (73.8) |  |
| Undecided | 150 | (21.5) | 33 | (26.8) | 117 | (20.6) |  |
| No | 49 | (7.0) | 16 | (13.0) | 32 | (5.6) |  |
| Desire to raise children |  |  |  |  |  |  | <.001 |
| Yes | 485 | (69.4) | 69 | (56.1) | 411 | (72.4) |  |
| Undecided | 146 | (20.9) | 28 | (22.8) | 116 | (20.4) |  |
| No | 67 | (9.6) | 25 | (20.3) | 41 | (7.2) |  |
| Unknown | 1 | (0.1) | 1 | (0.8) | 0 | (0.0) |  |
| Learning about child rearing |  |  |  |  |  |  | .321 |
| Yes | 528 | (75.5) | 89 | (72.4) | 435 | (76.6) |  |
| No | 168 | (24.0) | 34 | (27.6) | 133 | (23.4) |  |
| Unknown | 3 | (0.4) | 0 | (0.0) | 0 | (0.0) |  |
| Learning about the value of life |  |  |  |  |  |  | .609 |
| Yes | 680 | (97.3) | 119 | (96.7) | 557 | (98.1) |  |
| No | 14 | (2.0) | 3 | (2.4) | 10 | (1.8) |  |
| Unknown | 5 | (0.7) | 1 | (0.8) | 1 | (0.2) |  |

*Note.* The total analytic sample comprised 699 participants. Group comparisons by abuse experience were based on 691 participants because data on abuse experience were missing for eight participants. Group differences were examined using chi-square tests. Missing responses are shown as “Unknown”.

Exploratory factor analysis of the acceptability of corporal punishment (AACP) identified a three-factor structure (Table 3). Factor 1 consisted of items representing neglect and psychological maltreatment, Factor 2 represented harsh physically aggressive behaviors, and Factor 3 represented less severe aggressive behaviors. The three factors explained 31.2%, 8.09%, and 3.89% of the variance, respectively. Internal consistency was acceptable to good, with a Cronbach’s alpha coefficient of .884 for the total AACP scale. Cronbach’s alpha coefficients for the three subfactors were .843 for Factor 1, .861 for Factor 2, and .763 for Factor 3. Correlations among the three subfactors were all positive and statistically significant. Factor 1 was moderately correlated with Harsh physically aggressive behaviors (r = .576, p < .01) and Less severe aggressive behaviors (r = .469, p < .01), while Harsh physically aggressive behaviors was also moderately correlated with Less severe aggressive behaviors (r = .593, p < .01).

**Table 3.** Factor loadings of the acceptability of corporal punishment (AACP)

|  | Factor 1<br>*Neglect and psychological maltreatment* | Factor 2<br>*Harsh physically aggressive behaviors* | Factor 3<br>*Less severe aggressive behaviors* |
|---|---|---|---|
| Withholding bathing | .65 |  |  |
| Ignoring the child | .63 |  |  |
| Leaving child unclothed | .56 |  |  |
| Not changing underwear | .56 |  |  |
| Verbal threats | .52 |  |  |
| Withholding food | .49 |  |  |
| Favoring other sibling(s) | .48 |  |  |
| Leaving child unattended in a car | .47 |  |  |
| Preventing school attendance | .47 |  |  |
| Confinement in a room/closet | .46 |  |  |
| Putting child outside (e.g., balcony) | .45 |  |  |
| Leaving child home alone (for recreation) | .41 |  |  |
| Slapping the face |  | .78 |  |
| Kicking legs |  | .78 |  |
| Throwing objects |  | .63 |  |
| Slapping the head |  | .60 |  |
| Pinching |  | .56 |  |
| Forced haircut |  | .31 |  |
| Slapping the hands |  |  | .70 |
| Spanking (buttocks) |  |  | .70 |
| Harsh verbal scolding |  |  | .62 |
| Leaving a child alone while crying |  |  | .45 |
| Cronbach’s alpha coefficient | .843 | .861 | .763 |
| Proportion of variance explained (%) | 31.2 | 8.09 | 3.89 |

*Note.* Factors were extracted using the principal factor method and rotated using varimax rotation. Item labels are abbreviated for publication and do not reproduce the full original wording. The full set of items is available in the original source (Lee & Yasuyama, 2002).

Among the 699 participants, 123 (17.6%) reported at least one abuse experience. Of the five abuse categories, psychological abuse was the most commonly reported (13.3%), followed by physical abuse (8.4%) and witnessing domestic violence (8.3%). By contrast, sexual abuse (0.6%) and neglect (1.4%) were reported less frequently (Supplementary Table S1).

For the AACP items, harsh verbal scolding was the behavior most commonly judged to be acceptable (27.9%) (Table 4). This was followed by slapping a child’s hands (17.6%), leaving a child alone while crying (14.0%), and spanking a child’s buttocks (10.0%). In contrast, no participants endorsed not bathing a child or not changing a child’s underwear as acceptable. Taken together, these findings suggest that relatively mild aggressive or verbally harsh behaviors were more likely to be tolerated than severe physical or neglect-related acts. The total AACP score did not differ significantly between participants with and without abuse experiences based on the Mann–Whitney U test (p = .193).

**Table 4.** Responses to the acceptability of corporal punishment items (N = 699)

|  |  | n | % |
|---|---|---|---|
| 1 | Harsh verbal scolding | 195 | 27.9 |
| 2 | Spanking (buttocks) | 70 | 10.0 |
| 3 | Slapping the hands | 123 | 17.6 |
| 4 | Slapping the head | 40 | 5.7 |
| 5 | Slapping the face | 11 | 1.6 |
| 6 | Kicking legs | 12 | 1.7 |
| 7 | Pinching | 30 | 4.3 |
| 8 | Throwing objects | 7 | 1.0 |
| 9 | Forced haircut | 38 | 5.4 |
| 10 | Leaving a child alone while crying | 98 | 14.0 |
| 11 | Withholding food | 5 | 0.7 |
| 12 | Withholding bathing | 0 | 0.0 |
| 13 | Not changing underwear | 0 | 0.0 |
| 14 | Confinement in a room/closet | 11 | 1.6 |
| 15 | Putting child outside (e.g., balcony) | 21 | 3.0 |
| 16 | Leaving child home alone (for recreation) | 33 | 4.7 |
| 17 | Leaving child unattended in a car | 14 | 2.0 |
| 18 | Leaving child unclothed | 2 | 0.3 |
| 19 | Verbal threats | 29 | 4.1 |
| 20 | Ignoring the child | 15 | 2.1 |
| 21 | Favoring other sibling(s) | 2 | 0.3 |
| 22 | Preventing school attendance | 6 | 0.9 |

*Note.* Values indicate the number and percentage of participants who rated each behavior as either “acceptable as discipline” or “somewhat acceptable as discipline.” Item labels are abbreviated for publication and do not reproduce the full original wording. The full set of items is available in the original source (Lee & Yasuyama, 2002).

Subsequent analyses were restricted to participants without a history of abuse experience (n = 568), consistent with the aim of the present study. In this subgroup, the mean total AACP score was 82.8 (SD = 5.13). The mean scores for the three subfactors were 46.7 (SD = 2.21) for Factor 1 (Neglect and psychological maltreatment), 23.0 (SD = 1.90) for Factor 2 (Harsh physically aggressive behaviors), and 13.1 (SD = 2.25) for Factor 3 (Less severe aggressive behaviors).

In the univariate analyses, the total AACP score was significantly associated with sex, faculty, economic situation, experience of physical violence toward oneself, belief that corporal punishment is necessary for discipline, interest in child abuse, and gender role view (Supplementary Table S2). For the subfactors, Factor 1 (Neglect and psychological maltreatment) was significantly associated with economic situation, experience of physical violence toward oneself, belief that corporal punishment is necessary for discipline, and gender role view. Factor 2 (Harsh physically aggressive behaviors) was significantly associated with sex, number of siblings, living arrangement in childhood, belief that corporal punishment is necessary for discipline, interest in child abuse, and gender role view. Factor 3 (Less severe aggressive behaviors) was significantly associated with belief that corporal punishment is necessary for discipline and gender role view.

In the multivariable linear regression analyses, gender role view remained significantly associated with the total AACP score and with the two aggression-related subfactors, that is, Factor 2 (Harsh physically aggressive behaviors) and Factor 3 (Less severe aggressive behaviors), although its association with Factor 1 did not reach statistical significance (Table 5). Experience of physical violence toward oneself was significantly associated with the total AACP score, Factor 1, and Factor 3. In addition, learning about child rearing and being enrolled in a faculty related to health, welfare, and rehabilitation were significantly associated with Factor 3. Witnessing psychological violence toward others was negatively associated with Factor 3; given the coding of this variable, participants with more frequent witnessing experiences tended to have higher Factor 3 scores, indicating lower acceptance of less severe aggressive behaviors. In contrast, female sex was positively associated with Factor 2, suggesting lower acceptance of harsh physically aggressive behaviors among women.

**Table 5.** Factors associated with AACP and subfactor scores among those without abuse experience: multivariable linear regression analysis (N = 568)

| Variables | AACP |  |  |  | Factor 1<br>*Neglect and psychological maltreatment* |  |  |  | Factor 2<br>*Harsh physically aggressive behaviors* |  |  |  | Factor 3<br>*Less severe aggressive behaviors* |  |  |  |
|---|---|---|---|---|---|---|---|---|---|---|---|---|---|---|---|---|
|  | *B* <sup>b</sup> | *SE* <sup>c</sup> | *β* <sup>d</sup> | *p*-value | *B* <sup>b</sup> | *SE* <sup>c</sup> | *β* <sup>d</sup> | *p*-value | *B* <sup>b</sup> | *SE* <sup>c</sup> | *β* <sup>d</sup> | *p*-value | *B* <sup>b</sup> | *SE* <sup>c</sup> | *β* <sup>d</sup> | *p*-value |
| Sex<br>(ref. male) | 1.02 | 0.54 | 0.09 | .062 |  |  |  |  | 0.69 | 0.19 | 0.16 | <.001 |  |  |  |  |
| Living arrangement in childhood<br>(ref. with parents) | 0.98 | 0.57 | 0.08 | .087 |  |  |  |  | 0.36 | 0.21 | 0.08 | .086 |  |  |  |  |
| Gender role view<br>1. Strongly agree<br>2. Agree<br>3. Disagree<br>4. Strongly disagree | 1.40 | 0.40 | 0.16 | .001 | 0.33 | 0.17 | 0.09 | .052 | 0.56 | 0.15 | 0.17 | <.001 | 0.66 | 0.17 | 0.17 | <.001 |
| Experience of physical violence toward oneself<br>1. Often<br>2. Sometimes<br>3. Never | 1.49 | 0.66 | 0.11 | .024 | 0.84 | 0.26 | 0.15 | .001 |  |  |  |  | 0.64 | 0.28 | 0.11 | .022 |
| Witnessing psychological violence toward others<br>1. Often<br>2. Sometimes<br>3. Never | –0.87 | 0.49 | –0.09 | .073 |  |  |  |  |  |  |  |  | –0.44 | 0.21 | –0.10 | .038 |
| Learning about child rearing<br>(ref. No) | 1.05 | 0.56 | 0.08 | .061 |  |  |  |  | 0.37 | 0.20 | 0.08 | .069 | 0.49 | 0.25 | 0.09 | .049 |
| Health, Welfare, and Rehabilitation faculty<br>(ref. MDP<sup>a</sup>) |  |  |  |  |  |  |  |  |  |  |  |  | 0.46 | 0.21 | 0.10 | .031 |
| Interaction with children<br>1. Often<br>2. Once or a few times<br>3. Not at all |  |  |  |  |  |  |  |  |  |  |  |  | 0.33 | 0.18 | 0.08 | .072 |

*Note.* Variable reduction was applied.
<sup>a</sup>Medicine, Dentistry, and Pharmacology. <sup>b</sup>Unstandardized regression coefficient, <sup>c</sup>Standard error, <sup>d</sup>Standardized regression coefficient.

## 4. Discussion

This study examined factors associated with the acceptance of corporal punishment among university students without childhood maltreatment experiences. Gender-role attitudes, violence-related experiences during school years, learning about child rearing, and professional education were associated with the total AACP score or its subfactor scores. At the item level, acceptance was low for behaviors representing severe physical violence or neglect. In contrast, behaviors that may be more easily regarded as part of everyday discipline, such as harsh verbal scolding and slapping a child’s hands or buttocks, were relatively more accepted. This pattern was consistent with findings from a previous study of male university students (Kit et al., 2026). In addition, the factors associated with AACP scores differed partly across the subfactors. The following sections discuss these associated factors and their implications for future educational interventions.

### 4.1. Interpretation of associated factors

#### 4.1.1. Gender-role attitudes

In this study, participants with more traditional gender-role attitudes had lower total AACP, Factor 2, and Factor 3 scores, indicating greater acceptance of corporal punishment and related harmful behaviors. This finding is consistent with a previous study of male university students, which showed that students with more egalitarian gender-role attitudes were less likely to accept corporal punishment as a form of discipline (Kit et al., 2026).

Previous studies have reported similarities in gender-role attitudes between parents and children, suggesting that these attitudes are shaped and passed on through socialization within families (Chen et al., 2024; Marks et al., 2009). Traditional masculinity norms have also been reported to include an emphasis on social status and toughness (Thompson & Pleck, 1986). Taken together, these findings suggest that values related to gender roles, developed through family and social experiences, may contribute to the association between gender-role attitudes and acceptance of corporal punishment. However, the present study cannot explain the specific mechanisms underlying this association.

Regarding sex, female participants had higher Factor 2 scores, indicating lower acceptance of harsh physically aggressive behaviors. This finding is consistent with a previous study reporting that male university students were more likely than female students to justify corporal punishment (Rajter & Medvidović, 2025). However, the present study did not directly examine how sex differences in these attitudes develop. Therefore, we cannot determine whether the observed association reflects biological differences or differences in gender-related socialization and social norms.

#### 4.1.2. Violence-related experiences during school years

Regarding violence-related experiences during school years, it is noteworthy that experiencing physical violence and witnessing psychological violence were associated with AACP scores in different directions. Participants who had not experienced physical violence had higher AACP scores, suggesting that experiences of physical violence during school years may be associated with greater acceptance of corporal punishment and related harmful behaviors toward children. In previous studies, experience of corporal punishment in educational settings, such as schools and extracurricular sports activities, has been associated with more favorable attitudes toward corporal punishment later in life (Uchida et al., 2020). In the present study, experiences of physical violence from teachers or other adults may have been interpreted as part of “guidance” or “discipline” rather than as inappropriate violence. Such interpretations may, in turn, be associated with greater acceptance of corporal punishment and related harmful behaviors in parent–child relationships.

In contrast, witnessing psychological violence was negatively associated with Factor 3. Participants who had witnessed psychological violence were less likely to accept less severe aggressive behaviors. Factor 3 includes behaviors such as harsh verbal scolding and leaving a child alone while crying, which may appear less physically severe but still involve psychological pressure or rejection. The mechanism underlying this association cannot be determined from the present study. However, social cognitive theory suggests that observing the behavior of others can contribute to the development of beliefs about whether a behavior is appropriate or acceptable (Bussey & Bandura, 1999). The present findings may therefore suggest that witnessing psychological violence is related to how individuals recognize psychologically harmful interactions. This interpretation remains hypothetical, and future studies should examine the specific nature of the violence witnessed and how individuals understood or interpreted those experiences. Importantly, this interpretation does not suggest that witnessing psychological violence is a desirable experience. Educational interventions should provide opportunities to understand the harmful effects of psychologically damaging interactions without relying on actual experiences of violence or witnessing violence.

#### 4.1.3. Learning about child rearing and professional education

Learning about child rearing was associated with Factor 3. In this study, “learning about child rearing” included learning opportunities during elementary school, junior high school, high school, university, and other periods of life. These opportunities were not limited to professional education and may have included school classes, home economics education, and other learning experiences. Students with such learning experiences may have had more opportunities to consider children’s development, emotions, and caregiver–child interactions. This may have helped them recognize behaviors that are often regarded as “minor” or “common” forms of discipline as potentially harmful to children. This interpretation is consistent with previous studies on child-rearing and child-care education during adolescence. A Japanese study of junior and senior high school students found that child-care learning in home economics classes, including opportunities to interact with young children, increased students’ interest in children, knowledge of child development, and empathic responsiveness toward children (Okano et al., 2012). Similarly, a study of high school students in the United States found that knowledge of child development, experience interacting with children, and coursework on child development were associated with more favorable beliefs about understanding children’s feelings and development, supporting children’s learning and social-emotional development, and parenting knowledge and efficacy (O’Donnell Weber et al., 2023). Taken together, learning about child rearing may promote a better understanding of children’s development, emotions, and responsive caregiving. Such learning may help students recognize behaviors such as harsh verbal scolding, slapping a child’s hands or buttocks, or leaving a crying child alone as behaviors that may threaten children’s psychological safety or development, even when these behaviors appear relatively mild. Students enrolled in health, welfare, and rehabilitation-related faculties had higher Factor 3 scores than students enrolled in medical, dental, or pharmaceutical faculties. Differences in educational content across academic fields may partly explain this finding. Students in health, welfare, and rehabilitation-related fields may have more opportunities to learn about the dignity, rights, development, and living circumstances of children, families, and people receiving support. Such learning may be associated with greater awareness of behaviors that can easily be overlooked as part of everyday discipline. Although the academic field differs, a previous study of Japanese university students reported that students majoring in child care were less likely to accept parental corporal punishment (Kitano et al., 2018). In addition, a study examining the relationship between college education and beliefs about parental discipline found that upper-year students were less supportive of spanking and harsh discipline than first-year students, and that taking courses that included content related to discipline was also associated with lower endorsement of punitive discipline (Tompkins, 2026). These findings suggest that professional learning experiences related to children and discipline may be associated with attitudes toward corporal punishment and harsh discipline. However, caution is needed when interpreting the differences observed across academic fields as an effect of professional education itself. Students’ own values and interests, family background, and socioeconomic factors may have been related both to their choice of academic field and to their Factor 3 scores. Therefore, the observed association may reflect not only differences in educational content, but also differences in students’ backgrounds before entering university. Because the present study did not assess the specific educational content of each academic field or students’ pre-university backgrounds, future studies should take these factors into account.

### 4.2. Conceptual summary of factors associated with corporal punishment acceptability among young adults without childhood maltreatment experiences

The present findings showed that gender-role attitudes, violence-related experiences during school years, learning about child rearing, and professional education were associated with corporal punishment acceptability among university students without childhood maltreatment experiences. These associations also differed partly across the AACP subfactors. Based on these findings, we developed a conceptual summary that places these experiences, observations, and learning opportunities along an approximate developmental timeline (Figure 1). Although sex was associated with Factor 2, sex itself was not included in the figure because it is not a modifiable factor. In contrast, gender-role attitudes may be shaped through family, school, and other social experiences and may change through education. Therefore, gender-role attitudes, which were directly measured in this study, were included in the figure as a potentially modifiable factor.

**Figure 1.**
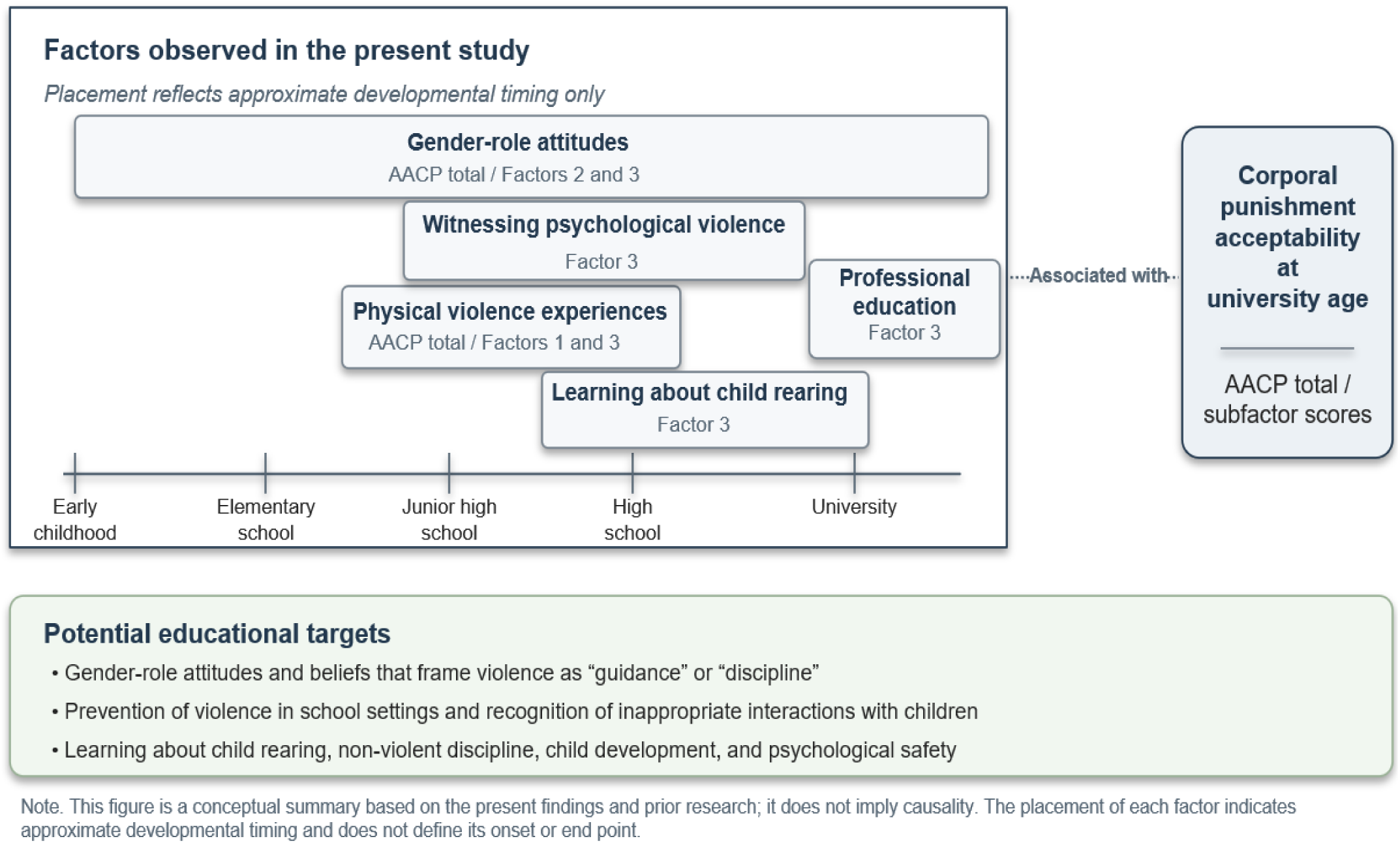
Conceptual summary of factors associated with corporal punishment acceptability among young adults without childhood maltreatment experiences.

Because this was a cross-sectional study, the figure does not represent causal relationships. Rather, it provides a conceptual summary based on the present findings and previous research. Future longitudinal and qualitative studies are needed to examine how experiences and learning at different developmental stages are related to the development and change of attitudes toward corporal punishment.

### 4.3. Implications for future interventions

The present findings suggest that education aimed at preventing the acceptance of corporal punishment should address more than the harmful effects of corporal punishment itself. It may also be important to address gender-role attitudes, beliefs that frame violence as “guidance” or “discipline,” children’s development and psychological harm, and nonviolent approaches to discipline. In particular, learning about child rearing and professional education were associated with Factor 3. This factor includes behaviors such as harsh verbal scolding, slapping a child’s hands or buttocks, and leaving a crying child alone.

These behaviors may not always be recognized as serious violence and may instead be accepted as ordinary forms of discipline. Therefore, learning opportunities related to child rearing and child development may benefit from addressing these behaviors directly and considering their possible effects on children’s psychological safety and development, together with nonviolent approaches to discipline.

Moreover, because attitudes toward corporal punishment were associated with experiences and learning occurring at different developmental stages, education should not be limited to a single intervention at university. Instead, developmentally appropriate and continuous educational approaches should be considered. Such a multidimensional approach is consistent with the INSPIRE framework (World Health Organization, 2016), which combines strategies related to norms and values, parent and caregiver support, and education and life skills.

### 4.4. Limitations

This study has several limitations. First, because this was a cross-sectional study, causal relationships between the explanatory variables and attitudes toward corporal punishment cannot be established. In particular, regarding professional education, students’ own values, interests, and family backgrounds may have been related to both their choice of academic field and their attitudes toward corporal punishment. Therefore, caution is needed when interpreting the differences between academic fields as an effect of professional education itself. In addition, the analysis did not account for differences in the content or extent of learning about child rearing. Therefore, we could not determine which types of learning experiences were associated with attitudes toward corporal punishment.

Second, violence experiences and witnessing violence were assessed using self-reported measures and may therefore have been affected by recall bias and social desirability bias. We also did not obtain detailed information about the nature or frequency of these experiences, the relationship with the person who used violence, or how participants interpreted the experiences. Furthermore, the items used to assess experiences of and exposure to violence during school years were developed specifically for this study, and their validity and reliability have not been fully established.

Third, childhood maltreatment history was based on self-report. Therefore, the group classified as having no childhood maltreatment experiences may have included participants who experienced inappropriate caregiving but did not recognize or remember it as maltreatment. In addition, five items were selected from the existing Japanese version of the ACE questionnaire, and their wording was simplified and partially modified to facilitate participants’ responses. Therefore, the validity and reliability of the five items used in the present study cannot be assumed to be equivalent to those of the original or Japanese versions.

Fourth, participants were recruited from a limited number of universities, and the response rate was low (10.2%). Therefore, caution is required when generalizing the findings, and selection bias cannot be excluded. In addition, sex was assessed using a limited set of response categories, and gender identity was not measured separately. Thus, findings related to sex should be interpreted with caution.

Fifth, the AACP subfactors were derived from exploratory factor analysis in the present study, and their reproducibility should be examined in other populations. Although the subfactors represent different types of behaviors, they all share the broader concept of how acceptable participants consider harmful behaviors toward children. Therefore, differences in associated factors across the subfactors should not be interpreted as indicating completely independent constructs.

Despite these limitations, this study examined factors associated with corporal punishment acceptability among young adults without childhood maltreatment experiences from several perspectives, including values, violence-related experiences, learning about child rearing, and professional education. It also showed that the associated factors differed partly across AACP subfactors. Future qualitative and longitudinal studies are needed to clarify how attitudes toward corporal punishment develop and change over time.

## 5. Conclusion

This study examined factors associated with corporal punishment acceptability among university students without childhood maltreatment experiences. Gender-role attitudes, violence-related experiences during school years, learning about child rearing, and professional education were associated with the total AACP score or its subfactor scores. The associated factors also differed partly across neglect and psychological maltreatment, harsh physically aggressive behaviors, and less severe aggressive behaviors.

These findings suggest that, even among young adults without childhood maltreatment experiences at home, attitudes toward corporal punishment may be related to gender-role values, violence-related experiences during school years, and learning about children and child rearing. The factors identified in this study provide a basis for understanding why some young adults without childhood maltreatment experiences may still accept corporal punishment and for considering educational approaches to prevent the development of such attitudes. Future interventions should go beyond simply communicating the harmful effects of corporal punishment and should consider developmentally appropriate and continuous education addressing beliefs that frame violence as “guidance” or “discipline,” children’s development and psychological harm, and nonviolent approaches to discipline.

## Ethics approval and consent to participate

This study was approved by the Ethics Committee of Kyoto Koka Women’s University (approval No. 23-15). Participants were provided with study information before beginning the online survey and indicated consent by selecting “I agree to participate.” Participation was voluntary.

## Consent for publication

Not applicable. No information identifying individual participants is included in this manuscript.

## Data availability

The individual-level data are not publicly available because they contain sensitive self-reported information about childhood maltreatment and violence. Aggregate findings are provided in the manuscript and Supplementary Material.

## Competing interests

The authors declare that they have no competing interests.

## Funding

This study was supported by JSPS KAKENHI Grant No. 21K11143. The funder had no role in the study design, data collection, analysis, interpretation, manuscript preparation, or decision to submit the article.

## Acknowledgements

We wish to thank the participants who responded to the questionnaire in this study.

## CRediT author contributions

Ayano Kit: Conceptualization, Methodology, Investigation, Data curation, Formal analysis, Funding acquisition, Project administration, Writing – original draft, Writing – review & editing.

Asuka Miyazaki: Conceptualization, Methodology, Investigation, Formal analysis, Writing – review & editing.

Naomi Kitano: Conceptualization, Methodology, Investigation, Writing – review & editing. Tetsuji Okawachi: Investigation, Writing – review & editing.

Andari Wuri Astuti: Writing – review & editing.

Sumihisa Honda: Conceptualization, Methodology, Investigation, Formal analysis, Writing – review & editing, Supervision.

## Declaration of generative AI and AI-assisted technologies in the manuscript preparation process

ChatGPT (OpenAI) was used to assist with English editing and manuscript organization. The authors reviewed and revised the content and take full responsibility for the manuscript.

## Notes

### Competing Interest Statement

The authors have declared no competing interest.

### Author Declarations

The Ethics Committee of Kyoto Koka Women's University granted ethical approval for this study (Approval No. 23-15).

