## Supplementary Material for "Corporal punishment acceptability among university students without childhood maltreatment experiences: Associated factors and subfactor-specific patterns"

This supplementary material provides additional information on the questionnaire items and statistical analyses used in the present study. Appendix A presents the study-specific questionnaire items, including their response options and coding for analysis. Appendix B presents the five adapted items used to assess childhood maltreatment experiences. Supplementary Table S1 summarizes childhood maltreatment experiences in the total sample, and Supplementary Table S2 presents the univariate analyses of AACP and subfactor scores among participants without childhood maltreatment experiences.

**Appendix A. Study-specific Questionnaire Items Used in the Analyses**

| Variable | Item used in the present study | Original response options | Coding used in the analyses |
|---|---|---|---|
| Interaction with children | Have you ever had opportunities to care for or play with young children, including younger siblings? Please include practicum or other hands-on experiences. | Often / Once or a few times / Not at all | Often / Once or a few times / Not at all |
| Having been taught that corporal punishment is necessary | Have you ever been taught by a school teacher, school staff member, or someone in an extracurricular educational setting (e.g., a cram school, extracurricular lesson, or after-school program) that "corporal punishment is necessary for a person's growth"? | Yes / No | Yes / No |

| | | | |
|---|---|---|---|
| Belief that corporal punishment is necessary for discipline | Do you agree or disagree with the statement, “Corporal punishment of children is necessary for discipline”? | Strongly agree / Agree / Disagree / Strongly disagree | Agree = Strongly agree + Agree; Disagree = Disagree + Strongly disagree |
| Interest in child abuse | How interested are you in child abuse? | Interested / Somewhat interested / Not very interested / Not interested | Yes = Interested + Somewhat interested; No = Not very interested + Not interested |
| Learning about child rearing | Have you ever learned about child rearing? (e.g., the joys and difficulties of raising children, household management, and parent–child affection) | Yes / No | Yes / No |
| Learning about the value of life | Have you ever attended a class about the value of life? (e.g., sexuality, the process from conception to birth, and the importance of life) | Yes / No | Yes / No |

Note. These study-specific items were developed for the present study and were administered in Japanese. The English wording shown here is a translation of the original Japanese questionnaire. For items originally assessed using four response categories, responses were dichotomized for analysis as indicated in the table.

**Appendix B. Adapted Items Used to Assess Childhood Maltreatment Experiences**

| Domain | Item used in the present study | Response |
|---|---|---|
| Physical abuse | Before the age of 18, did your father, mother, or another adult living with you repeatedly use physical violence against you (e.g., hitting or kicking)? | Yes / No |

| | | |
|---|---|---|
| Psychological/emotional abuse | Before the age of 18, did your father, mother, or another adult living with you repeatedly use psychological violence against you (e.g., hurtful or abusive language)? | Yes / No |
| Sexual abuse | Before the age of 18, did you experience any sexual abuse by your father, mother, or another adult living with you? | Yes / No |
| Emotional neglect | Before the age of 18, did you feel that no one in your family cared about you or that you lacked a sense of connection with your family? | Yes / No |
| Exposure to family violence | Before the age of 18, was any member of your family frequently pushed, grabbed, slapped, or had objects thrown at them? | Yes / No |

Note. These items were adapted from the ACE questionnaire (Felitti et al., 1998) and the Japanese version developed by Tsuboi (2014). The wording was simplified and partially modified for the present study.

**Supplementary Table S1. Childhood Maltreatment Experiences in the Total Sample (N = 699)**

| | Yes | | No | | Missing | |
|---|---|---|---|---|---|---|
| | n | % | n | % | n | % |
| Any abuse experience | 123 | 17.6 | 568 | 81.3 | 8 | 1.1 |
| Physical abuse | 59 | 8.4 | 635 | 90.8 | 5 | 0.7 |
| Psychological abuse | 93 | 13.3 | 601 | 86.0 | 5 | 0.7 |
| Sexual abuse | 4 | 0.6 | 689 | 98.6 | 6 | 0.9 |
| Exposure to domestic violence | 58 | 8.3 | 635 | 90.8 | 6 | 0.9 |
| Neglect | 10 | 1.4 | 684 | 97.9 | 5 | 0.7 |

Note. Percentages were calculated using the total sample (N = 699) as the denominator. Missing values are shown separately for each item.

**Supplementary Table S2. Univariate Associations of Participant Characteristics with AACP and Subfactor Scores Among Students without Childhood Maltreatment Experiences (N = 568)**

| | AACP | | Factor 1 *Neglect and psychological maltreatment* | | Factor 2 *Harsh physically aggressive behaviors* | | Factor 3 *Less severe aggressive behaviors* | |
|---|---|---|---|---|---|---|---|---|
| | Mean | p-value | Mean | p-value | Mean | p-value | Mean | p-value |
| Sex | | .025[a] | | .185[a] | | <.001[a] | | .067[a] |
| Female | 83.2 | | 46.8 | | 23.2 | | 13.2 | |
| Male | 81.7 | | 46.5 | | 22.5 | | 12.8 | |
| University grade | | .387[b] | | .548[b] | | .753[b] | | .483[b] |
| 1st | 83.1 | | 46.8 | | 23.1 | | 13.2 | |
| 2nd | 82.7 | | 46.6 | | 23.0 | | 13.2 | |
| 3rd | 82.2 | | 46.6 | | 22.8 | | 12.8 | |
| 4th | 83.0 | | 46.7 | | 23.1 | | 13.2 | |
| 5th | 81.3 | | 45.9 | | 22.8 | | 12.5 | |
| 6th | 82.9 | | 46.8 | | 23.1 | | 13.0 | |
| Place of birth | | .917[b] | | .556[b] | | .907[b] | | .545[b] |
| Kanto area | 83.4 | | 46.4 | | 23.4 | | 13.7 | |
| Kansai area | 82.6 | | 46.7 | | 22.9 | | 13.0 | |
| Kyushu area | 82.8 | | 46.6 | | 23.0 | | 13.2 | |
| Others | 83.1 | | 46.9 | | 23.1 | | 13.0 | |
| Faculty | | .049[b] | | .069[b] | | .202[b] | | .137[b] |
| Medicine, Dentistry and Pharmacology | 82.0 | | 46.4 | | 22.8 | | 12.8 | |
| Health, Welfare, and Rehabilitation | 83.3 | | 46.9 | | 23.1 | | 13.3 | |
| Child Education | 83.5 | | 47.0 | | 23.3 | | 13.1 | |
| Others | 83.4 | | 46.9 | | 23.2 | | 13.3 | |
| Economic situation | | .049[b] | | .028[b] | | .153[b] | | .230[b] |
| Poor | 81.5 | | 45.9 | | 23.0 | | 12.6 | |
| Normal | 83.2 | | 46.9 | | 23.1 | | 13.2 | |
| Rich | 82.4 | | 46.6 | | 22.8 | | 13.0 | |

| | | | | | | | | |
|---|---|---|---|---|---|---|---|---|
| Current living status | | .488[b] | | .972[b] | | .144[b] | | .370[b] |
| Alone | 83.0 | | 46.7 | | 23.1 | | 13.3 | |
| With parents | 82.8 | | 46.7 | | 23.0 | | 13.1 | |
| Others | 81.4 | | 46.3 | | 22.6 | | 12.5 | |
| Number of siblings (including the respondent) | | | | | | | | |
| | | .151[b] | | .679[b] | | .037[b] | | .246[b] |
| 1 (only child) | 82.8 | | 46.5 | | 23.0 | | 13.3 | |
| 2 | 82.4 | | 46.6 | | 22.9 | | 13.0 | |
| ≥3 | 83.3 | | 46.8 | | 23.3 | | 13.2 | |
| Living arrangement in childhood | | .085[a] | | .258[a] | | .017[a] | | .094[a] |
| With parents | 82.5 | | 46.6 | | 22.9 | | 13.0 | |
| Others | 83.8 | | 46.9 | | 23.4 | | 13.4 | |
| Partner status | | .217[a] | | .628[a] | | .796[a] | | .129[a] |
| Having | 82.6 | | 46.7 | | 23.1 | | 12.8 | |
| Not having | 82.8 | | 46.6 | | 23.0 | | 13.2 | |
| Mother's final education | | .329[b] | | .206[b] | | .621[b] | | .316[b] |
| Under high school | 83.5 | | 46.9 | | 23.2 | | 13.4 | |
| Above high school | 82.6 | | 46.6 | | 23.0 | | 13.0 | |
| Others | 82.4 | | 46.4 | | 22.9 | | 13.2 | |
| Father's final education | | .910[b] | | .742[b] | | .767[b] | | .852[b] |
| Under high school | 83.2 | | 46.9 | | 23.2 | | 13.1 | |
| Above high school | 82.6 | | 46.6 | | 23.0 | | 13.1 | |
| Others | 83.1 | | 46.9 | | 23.1 | | 13.3 | |
| Interaction with the community | | .487[a] | | .246[a] | | .100[a] | | .721[a] |
| Yes | 82.6 | | 46.5 | | 22.9 | | 13.1 | |
| No | 82.8 | | 46.7 | | 23.1 | | 13.1 | |
| Interaction with children | | .999[b] | | .985[b] | | .534[b] | | .829[b] |
| Often | 82.9 | | 46.8 | | 23.1 | | 13.0 | |
| Once or a few times | 82.6 | | 46.6 | | 22.9 | | 13.2 | |
| Not at all | 82.9 | | 46.4 | | 23.2 | | 13.2 | |
| Experience of physical violence toward oneself | | | | | | | | |
| | | .038[b] | | .005[b] | | .534[b] | | .088[b] |
| Often | 78.8 | | 45.0 | | 22.9 | | 10.9 | |
| Sometimes | 82.1 | | 46.2 | | 22.8 | | 13.1 | |
| Never | 82.9 | | 46.8 | | 23.1 | | 13.1 | |

| | | | | | | | | |
|---|---|---|---|---|---|---|---|---|
| Experience of psychological violence toward oneself | | | | | | | | |
| | | .207[b] | | .105[b] | | .466[b] | | .182[b] |
| Often | 80.3 | | 45.8 | | 22.9 | | 11.7 | |
| Sometimes | 82.5 | | 46.4 | | 22.8 | | 13.3 | |
| Never | 82.9 | | 46.7 | | 23.1 | | 13.1 | |
| Witnessing physical violence toward others | | | | | | | | |
| | | .481[b] | | .061[b] | | .181[b] | | .488[b] |
| Often | 81.4 | | 46.5 | | 22.8 | | 12.1 | |
| Sometimes | 82.6 | | 46.3 | | 23.1 | | 13.2 | |
| Never | 82.9 | | 46.8 | | 23.0 | | 13.1 | |
| Witnessing psychological violence toward others | | | | | | | | |
| | | .564[b] | | .544[b] | | .343[b] | | .314[b] |
| Often | 83.7 | | 46.8 | | 23.6 | | 13.3 | |
| Sometimes | 82.9 | | 46.6 | | 23.1 | | 13.2 | |
| Never | 82.7 | | 46.7 | | 23.0 | | 13.0 | |
| Having been taught that corporal punishment is necessary | | | | | | | | |
| | | .534[a] | | .273[a] | | .975[a] | | .914[a] |
| Yes | 82.7 | | 46.8 | | 22.8 | | 13.1 | |
| No | 82.8 | | 46.7 | | 23.0 | | 13.1 | |
| Belief that corporal punishment is necessary for discipline | | | | | | | | |
| | | <.001[a] | | .010[a] | | <.001[a] | | <.001[a] |
| Agree | 76.8 | | 45.7 | | 20.7 | | 10.4 | |
| Disagree | 83.1 | | 46.7 | | 23.2 | | 13.3 | |
| Interest in child abuse | | .014[a] | | .082[a] | | .004[a] | | .068[a] |
| Yes | 83.0 | | 46.7 | | 23.1 | | 13.2 | |
| No | 81.0 | | 46.1 | | 22.2 | | 12.5 | |
| Gender role view | | <.001[a] | | .026[a] | | <.001[a] | | <.001[a] |
| Agree | 80.4 | | 46.1 | | 22.1 | | 12.2 | |
| Disagree | 83.3 | | 46.8 | | 23.2 | | 13.3 | |
| Desire for marriage | | .615[b] | | .793[b] | | .718[b] | | .505[b] |
| Yes | 82.7 | | 46.7 | | 23.0 | | 13.0 | |
| Undecided | 82.8 | | 46.6 | | 23.0 | | 13.2 | |
| No | 83.4 | | 46.7 | | 23.3 | | 13.4 | |

| | | | | | | | | |
|---|---|---|---|---|---|---|---|---|
| Desire to raise children | | .906[b] | | .809[b] | | .655[b] | | .958[b] |
| Yes | 82.7 | | 46.7 | | 23.0 | | 13.1 | |
| Undecided | 83.0 | | 46.6 | | 23.2 | | 13.2 | |
| No | 82.5 | | 46.5 | | 23.0 | | 13.0 | |
| Learning about child rearing | | .148[a] | | .295[a] | | .483[a] | | .144[a] |
| Yes | 83.0 | | 46.7 | | 23.1 | | 13.2 | |
| No | 82.1 | | 46.6 | | 22.8 | | 12.8 | |
| Learning about the value of life | | .147[a] | | .498[a] | | .051[a] | | .155[a] |
| Yes | 82.8 | | 46.7 | | 23.1 | | 13.1 | |
| No | 81.6 | | 47.3 | | 22.0 | | 12.3 | |

*Note.* AACP = Attitudes toward the Acceptability of Corporal Punishment. Higher scores indicate lower acceptance of corporal punishment and related harmful behaviors.
[a]Mann–Whitney U test. [b]Kruskal–Wallis test.
